# Impact of HIV Infection on COVID-19 Outcomes Among Hospitalized Adults in the U.S.

**DOI:** 10.1101/2021.04.05.21254938

**Authors:** Matthew S. Durstenfeld, Kaiwen Sun, Yifei Ma, Fatima Rodriguez, Eric A. Secemsky, Rushi V. Parikh, Priscilla Y. Hsue

## Abstract

**Background:** Whether HIV infection is associated with differences in clinical outcomes among people hospitalized with COVID-19 is uncertain.

**Objective:** To evaluate the impact of HIV infection on COVID-19 outcomes among hospitalized patients.

**Methods:** Using the American Heart Association’s COVID-19 Cardiovascular Disease registry, we used hierarchical mixed effects models to assess the association of HIV with in-hospital mortality accounting for patient demographics and comorbidities and clustering by hospital. Secondary outcomes included major adverse cardiac events (MACE), severity of illness, and length of stay (LOS).

**Results:** The registry included 21,528 hospitalization records of people with confirmed COVID-19 from 107 hospitals in 2020, including 220 people living with HIV (PLWH). PLWH were younger (56.0+/-13.0 versus 61.3+/-17.9 years old) and more likely to be male (72.3% vs 52.7%), Non-Hispanic Black (51.4% vs 25.4%), on Medicaid (44.5% vs 24.5), and active tobacco users (12.7% versus 6.5%).

Of the study population, 36 PLWH (16.4%) had in-hospital mortality compared with 3,290 (15.4%) without HIV (Risk ratio 1.06, 95%CI 0.79-1.43; risk difference 0.9%, 95%CI −4.2 to 6.1%; p=0.71). After adjustment for age, sex, race, and insurance, HIV was not associated with in-hospital mortality (aOR 1.13; 95%CI 0.77-1.6; p 0.54) even after adding body mass index and comorbidities (aOR 1.15; 95%CI 0.78-1.70; p=0.48). HIV was not associated with MACE (aOR 0.99, 95%CI 0.69-1.44, p=0.91), severity of illness (aOR 0.96, 95%CI 0.62-1.50, p=0.86), or LOS (aOR 1.03; 95% CI 0.76-1.66, p=0.21).

**Conclusion:** HIV was not associated with adverse outcomes of COVID-19 including in-hospital mortality, MACE, or severity of illness.

**Condensed Abstract:** We studied 21,528 patients hospitalized with COVID-19 at 107 hospitals in AHA’s COVID-19 registry to examine the association between HIV and COVID-19 outcomes. More patients with HIV were younger, male, non-Hispanic Black, on Medicaid and current smokers. HIV was not associated with worse COVID-19 in-hospital mortality (Risk ratio 1.06, 95%CI 0.79-1.43; p=0.71) even after adjustment (aOR 1.15; 95%CI 0.78-1.70; p=0.48). HIV was also not associated with MACE (aOR 0.99, 95%CI 0.69-1.44, p=0.91) or severity of illness (aOR 0.96, 95%CI 0.62-1.50, p=0.86. Our findings do not support that HIV is a major risk factor for adverse COVID-19 outcomes.

## Introduction

There are limited data regarding risk of mortality among people living with human immunodeficiency virus (HIV; PLWH) hospitalized with COVID-19 (1). The prior severe acute respiratory syndrome epidemic suggested that people living with HIV/AIDS exposed to SARS-CoV-1 were not suspectable to infection with that coronavirus despite exposure (2). There is a case report of a patient with HIV/AIDs infected with EMC/2012, the coronavirus responsible for the Middle East Respiratory Syndrome epidemic which has a 60% mortality rate, who survived (3). Whether the experiences of PLWH with these prior coronaviridae are relevant to the present COVID-19 pandemic is uncertain.

A complex interplay of factors associated with HIV infection and treatment may alter clinical sequelae of COVID-19 among PLWH. First, treatment with antiretroviral therapy may decrease risk of severe disease or mortality after exposure to SARS-CoV-2, as protease inhibitors have been investigated as a possible therapy for COVID-19 (4). Second, if severe COVID-19 disease is related to excessive immune activation, immunosuppression that is present among treated and suppressed PLWH may alter the risk of severe disease or mortality in a way that might be protective. Alternatively, with excess cardiovascular risk due to traditional cardiovascular risk factors and heightened chronic inflammation/immune activation (5), PLWH may be more susceptible to cardiac injury or myocarditis from SARS-CoV-2 infection that results in higher mortality, cardiovascular complications, or post-acute sequelae of COVID-19 compared to non-HIV infected individuals. Prior work has demonstrated that PLWH with COVID-19 have higher prevalence of hypertension, diabetes, chronic kidney disease, and smoking compared to those with COVID-19 without HIV (6,7). Similar to the general population, traditional cardiovascular risk factors are associated with adverse COVID-19 outcomes among PLWH (6,7). Thus, SARS-CoV-2 infection may present with different disease severity and result in different clinical outcomes among PLWH (8).

Several studies have found higher risks of hospitalization (6,9) and mortality among PLWH (6,7) with COVID-19, but most case series of hospitalized patients suggest that the acute course of PLWH is similar to people without HIV (10-15). In contrast, the largest study of 122 PLWH hospitalized with COVID-19 in the United Kingdom found a higher adjusted risk of 28-day mortality (16). With these conflicting data, whether coinfection with HIV portends worse outcomes among hospitalized patients with COVID-19 remains uncertain (17). Therefore, we used the American Heart Association’s COVID-19 CVD Registry powered by Get with the Guidelines to study the impact of HIV infection with COVID-19 outcomes among a large sample of hospitalized patients in the United States.

## Methods

### Data Source/Study Population

The Get With The Guidelines® programs are provided by the American Heart Association (AHA). As described previously, the AHA COVID-19 Cardiovascular Disease Registry is a quality improvement program that began in April 2020 and as of December 2020 included 21,528 hospitalization records from 107 participating hospitals, including a mix of academic and community hospitals from across the United States (18). Approximately 2000 variables are collected, including hospital characteristics, demographics, medical history, symptoms, vital signs, lab tests, and clinical outcomes. Records are uploaded by participating hospitals and a deidentified dataset is provided to investigators after review by a scientific review committee. The dataset included for this study includes records entered through December 2020.

### Inclusion/Exclusion Criteria

Participating hospitals were instructed to upload data on all adult patients who had been hospitalized with confirmed COVID-19 and who had been discharged at the time of registry data upload. We included all patients uploaded to the registry in this analysis.

### Explanatory Variables

The primary predictor was HIV status as defined by the medical record. Data regarding antiretroviral therapy, viral load, CD4 counts, and AIDS status were not included in the database. Other explanatory variables included age, sex, race, insurance, body mass index (BMI), other past medical history including hypertension, dyslipidemia, diabetes, smoking, chronic kidney disease, chronic lung disease, and time from symptom onset to admission. Insurance status was classified as Medicare, Medicaid, commercial, self/uninsured, and other/unable to determine. We also report values for initial laboratory tests upon admission to the hospital.

### Outcomes

The primary outcome was in-hospital mortality, defined by a discharge disposition of “expired.” The first secondary outcome was a composite endpoint of major adverse cardiovascular outcomes (MACE) defined by in-hospital mortality, myocardial infarction, stroke, incident heart failure or myocarditis, and cardiogenic shock (19). The next secondary outcome was the AHA COVID-19 Ordinal Scale defined as died, cardiac arrest, mechanical ventilation and shock requiring mechanical circulatory support, mechanical ventilation with shock requiring vasopressors/inotropes, mechanical ventilation only, and hospitalization only (19). We also report components of MACE outcomes, mechanical ventilation, management in intensive care unit, and venous thromboembolism including deep venous thrombosis and pulmonary embolism. The next outcome was length of stay among survivors. Because PLWH were initially excluded from some COVID-19 clinical trials (20), the final secondary outcome was participation in a COVID-19 clinical trial.

### Missing Data

There were few missing data among our included covariates with exception of BMI, which was missing in 2,422 people (18 with HIV). We imputed missing values using multiple imputation with a random forest algorithm (MissRanger) based on the height, weight, age, sex, ZIP code, hospital, and history of diabetes. No other variables included in our models had a significant proportion of missing values.

### Statistical Analysis

To characterize the study population stratified by HIV status, we report numbers and proportions for categorical variables and means with standard deviations for continuous variables. We used chi-squared test or Fisher’s exact test for categorical variables, Student’s t-test for normally distributed continuous variables, Mann-Whitney U test for non-normally distributed continuous variables, and Kruskal-Wallis test for ordinal variables. We used hierarchical logistic regression mixed-effects models with hospital modeled as random effects and HIV and other covariates as fixed effects to examine associations of HIV status with our primary and secondary outcomes with both unadjusted models and models adjusted for demographics and past medical history and account for clustering by hospital. For the ordinal scale we used a hierarchical multinomial logistic regression mixed-effects model for adjusted analyses. We included models adjusted only for age, sex, race, and insurance in case comorbidities are mediators and models that are adjusted for age, sex, race, body mass index, and comorbidities to estimate the conditional effect of HIV independent of those potential confounders. P values for hierarchical models were generated by comparing the hierarchical model with HIV included with a similar model without HIV included using ANOVA. Wald estimates of confidence intervals were used for hierarchical models. P values of <0.05 were considered significant for the primary and secondary outcomes.

### Prespecified exploratory analyses

As an alternative approach to address confounding due to differences between those with and without HIV, we conducted a pre-specified exploratory propensity-matched analysis. We used a 3:1 control to case nearest neighbor matching algorithm (MatchIt) including the same variables as our adjusted model, including facility and the imputed BMI value for those missing BMI.

The American Heart Association Precision Medicine Platform (https://precision.heart.org/) was used for data analysis using R version 3.6.0 and SAS version 3.8. IQVIA (Parsippany, New Jersey) serves as the data collection and coordination center. Institutional Review Board approval was granted by the University of California, San Francisco.

## Results

Among 21,528 hospitalizations for COVID-19 at 107 hospitals, the mean age was 61.2+/-17.9 years and 9,884 (45.9%) were female. The population included 25.4% Hispanic/Latinx, 25.6% Non-Hispanic Black, 0.5% Native American, 4.0% Asian, 0.4% Pacific Islander, and 38.0% Non-Hispanic White. Of this cohort, there were 220 PLWH (1.0%), with characteristics described in detail in Table 1. PLWH were younger (56.0+/-13.0 versus 61.3+/-17.9 years old), more likely to be male (72.3% vs 52.7%), and more likely to be Non-Hispanic Black (51.4% vs 25.4%). PLWH were more likely to be on Medicaid (44.5% vs 24.5%) and less likely to be on Medicare (20.9% vs 30.3%). Time from symptom onset to admission was not different by HIV status (mean 6.2 vs 6.0; median 5 days for both). PLWH had lower mean BMI (29.4 versus 30.8 kg/m^2^) and were more likely to be active tobacco users (12.7% versus 6.5%). Laboratory findings are shown in Table 1: PLWH had lower serum creatinine (p=0.02) and higher d-dimer (p=0.01) compared to people without HIV, but no other significant differences. Despite having younger age, PLWH did not have significant differences in other comorbidities (Appendix: Supplementary Table A).

**Table 1:** Demographics and Baseline Characteristics by HIV status. Baseline demographics, body mass index, time from symptom onset to admission, and admission laboratory testing. *Missing <5%. °Missing 5-19%. ^^^ Missing 20-50%. # Missing 60-85%

|  | HIV positive<br>(n=220) | HIV negative<br>(n=21,308) |
| --- | --- | --- |
| Age | 56.0 ± 13.0 | 62.3 ± 17.9 |
| Female | 61 (27.7%) | 9,823 (46.1%) |
| Ethnicity/Race |  |  |
| Hispanic/Latinx | 60 (27.3%) | 5,418 (25.4%) |
| Non-Hispanic Black | 113 (51.4%) | 5,418 (25.4%) |
| Native American | 0 | 106 (0.5%) |
| Asian | 1 (0.5%) | 856 (4.0%) |
| Pacific Islander | 0 | 78 (0.4%) |
| Non-Hispanic White | 26 (11.8%) | 8,176 (38.4%) |
| Other/Unable to Determine | 20 (9.1%) | 1,256 (5.9%) |
| Insurance |  |  |
| Medicare | 46 (20.9%) | 6,448 (30.3%) |
| Medicaid | 98 (44.5%) | 5,228 (24.5%) |
| Commercial | 56 (25.5%) | 6,638 (31.2%) |
| Self/Uninsured | 13 (5.9%) | 2,077 (9.7%) |
| Other/Unable to Determine | 7 (3.2%) | 917 (4.3%) |
| Body Mass Index, kg/m<sup>2</sup> ° | 29.4 ± 7.9 | 30.8 ± 8.5 |
| Past Medical History |  |  |
| Hypertension | 125 (56.8%) | 12,548 (58.9%) |
| Dyslipidemia | 69 (31.4%) | 7,354 (34.5%) |
| Diabetes | 88 (40.0%) | 7,526 (35.3%) |
| Active Tobacco Use | 28 (12.7%) | 1,378 (6.5%) |
| Chronic Kidney Disease | 35 (15.9%) | 2,749 (12.9%) |
| Chronic Lung Disease | 48 (21.8%) | 3,967 (18.6%) |
| Symptom Onset to Admission (days) ^ | 5.0 (IQR 2-9) | 5.0 (IQR 2-8) |
| Initial Laboratory Testing |  |  |
| WBC Count, k/uL * | 6.70 (IQR 4.95-8.70) | 7.00 (IQR 5.10-9.70) |
| Platelet Count, k/uL * | 204 (IQR 143-260) | 204 (IQR 167-267) |
| Serum Creatinine, mg/dl * | 1.00 (IQR 0.94-1.64) | 1.13 (IQR 0.80-1.47) |
| C reactive protein, mg/L ^ | 64 (IQR 24-100) | 57 (IQR 14-111) |
| Ferritin, ng/ml ^ | 484 (IQR 273-1013) | 563 (IQR 239-1157) |
| Troponin, ng/L ^ | 10 (IQR 0-50) | 11 (IQR 0-50) |
| BNP, pg/mL # | 34 (IQR 19-94) | 64 (IQR 23-223) |
| NT-Pro-BNP, pg/mL # | 299 (IQR 61-1753) | 217 (IQR 75-1607) |
| D-Dimer, ng FEU/mL # | 850 (IQR 300-1231) | 600 (IQR 410-1660) |

Of the study population, 36 PLWH (16.4%) had the primary outcome of in-hospital mortality compared with 3,290 (15.4%) patients without HIV (Table 2). In hospital-mortality was not significantly different by HIV status in unadjusted analysis (Risk ratio 1.06, 95%CI 0.79-1.43; risk difference 0.9%, 95%CI −4.2 to 6.1%; p=0.71; Central Illustration).

**Table 2:** Crude Outcomes by HIV status. Major adverse cardiovascular outcomes (MACE) is defined as in-hospital mortality, myocardial infarction, stroke, incident heart failure or myocarditis, and cardiogenic shock. Totals of MACE do not add up to 100% as each patient was only counted once for MACE even with multiple MACE events. Mech=mechanical.

|  | HIV positive<br>(n=220) | HIV negative<br>(n=21,308) |
| --- | --- | --- |
| **In-Hospital Mortality** | 36 (16.4%) | 3,290 (15.4%) |
| **Major Adverse Cardiac Event** | 40 (18.2%) | 4,130 (19.4%) |
| Died | 36 (16.4%) | 3,290 (15.4%) |
| Myocardial Infarction | 4 (1.8%) | 651 (3.1%) |
| Stroke | 2 (0.9%) | 310 (1.5%) |
| Incident Heart Failure | 0 | 391 (1.8%) |
| Myocarditis | 2 (0.9%) | 66 (0.3%) |
| Cardiogenic Shock | 1 (0.5%) | 136 (0.6%) |
| **AHA COVID-19 Ordinal Scale** |  |  |
| Died | 36 (16.4%) | 3,290 (15.4%) |
| Cardiac Arrest | 0 | 93 (0.4%) |
| Mech. Ventilation + Mech. Circulatory Support | 0 | 51 (0.2%) |
| Mech. Ventilation + Vasopressors/Inotropes | 12 (5.5%) | 775 (3.6%) |
| Mechanical Ventilation Only | 6 (2.7%) | 1,188 (5.6%) |
| Hospitalization Only | 166 (75.5%) | 15,911 (74.7%) |
| **Venous Thromboembolism** |  |  |
| Deep Venous Thrombosis | 6 (2.7%) | 502 (2.4%) |
| Pulmonary Embolism | 3 (1.4%) | 382 (1.8%) |
| **Enrolled in COVID-19 Clinical Trial** | 19 (8.6%) | 1962 (9.2%) |
| **Length of Stay among Survivors, median** | 6 (IQR 4-11) | 6 (IQR 4-11) |

**Central Illustration:**
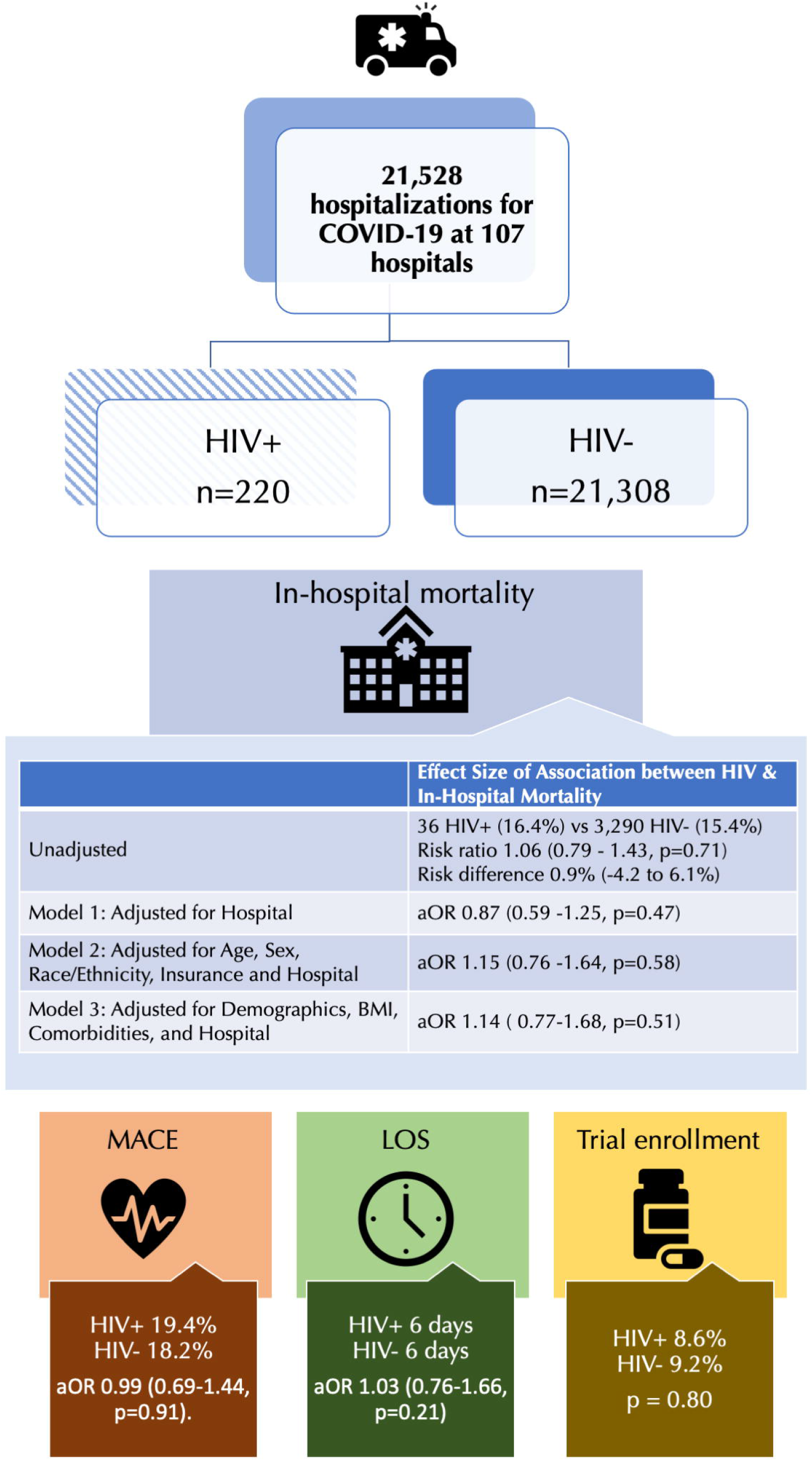
Association of HIV with COVID-19 Outcomes among Hospitalized Patients in the American Heart Association COVID-19 CVD Registry (n=21,528) HIV was not found to have a significant association with in-hospital mortality among people hospitalized with COVID-19 in unadjusted analysis (Risk ratio 1.06, 95%CI 0.79-1.43; p=0.71) or after adjustment (aOR 1.15; 95%CI 0.78-1.70; p=0.48). HIV was also not found to have an association with MACE (aOR 0.99, 95%CI 0.69-1.44, p=0.91) or severity of illness (aOR 0.96, 95%CI 0.62-1.50, p=0.86. or adjusted analyses. aOR=adjusted odds ratio; MACE=major adverse cardiac events; LOS=Length of Stay.

As shown in Table 3, in unadjusted hierarchical regression models to account for clustering by hospital, HIV was not associated with in-hospital mortality (Model 1: OR 0.87; 95% CI 0.59-1.25; P 0.47). After adjustment for age, sex, race, and insurance, HIV remained unassociated with in-hospital mortality (Model 2: aOR 1.13 95%CI 0.77-1.6; p 0.54). This relationship persisted after adding body mass index and comorbidities (Model 3: aOR 1.15; 95%CI 0.78-1.70; p=0.48).

**Table 3.** Adjusted Odds Ratios for Conditional Association of HIV with In-Hospital Mortality. HIV was not associated with in-hospital mortality in hierarchical mixed effects logistic regression analysis to account for clustering by hospital. Model 1 includes HIV (fixed effect) and hospital (random effects); Model 2 is adjusted for Age, Sex, Race, and Insurance; Model 3 is adjusted for demographics as in Model 2, body mass index, and past medical history. Confidence intervals are estimated via the Wald estimation and p-values are calculated by ANOVA compared to models excluding HIV.

|  | Odds Ratio | 95% CI | P value |
| --- | --- | --- | --- |
| Model 1 | 0.87 | 0.59-1.25 | 0.47 |
| Model 2 | 1.15 | 0.76-1.64 | 0.58 |
| Model 3 | 1.14 | 0.77-1.68 | 0.51 |

Among the entire population, 4,170 individuals (19.4%) had MACE which included 3,326 deaths, 655 with myocardial infarction, 312 with stroke, 391 with incident heart failure, 66 with myocarditis, and 136 with cardiogenic shock. Among PLWH, there were 40 MACE (36 deaths, 4 with myocardial infarction, 2 with stroke, 0 with incident heart failure, 2 with myocarditis, and 1 with cardiogenic shock; Table 2). Accounting for clustering by hospital and patient factors, HIV was not associated with increased risk of major adverse cardiac events (aOR 0.99; 95%CI 0.69-1.44; p=0.91). Patients with and without HIV also had similar incidence of deep venous thrombosis and pulmonary embolism (Table 2).

Among those with HIV, 47 (21.4%) required endotracheal intubation and mechanical ventilation compared to 4,152 (19.3%) without HIV (aOR 1.00; 95%CI. 0.71-1.40; p=1.00). Similarly, 59 PLWH (26.8%) compared to 6,604 without HIV (30.7%) were managed in an intensive care unit (aOR 0.90; 95%CI 0.65-1.23; p=0.49). Among those hospitalized with COVID-19, HIV was not associated with increased severity of illness as assessed by the American Heart Association’s COVID Ordinal Scale (Figure 1, p=0.93). In adjusted models, HIV was not associated with increased severity of illness (aOR 0.96; 95%CI 0.62-1.50; p=0.86).

**Figure 1:**
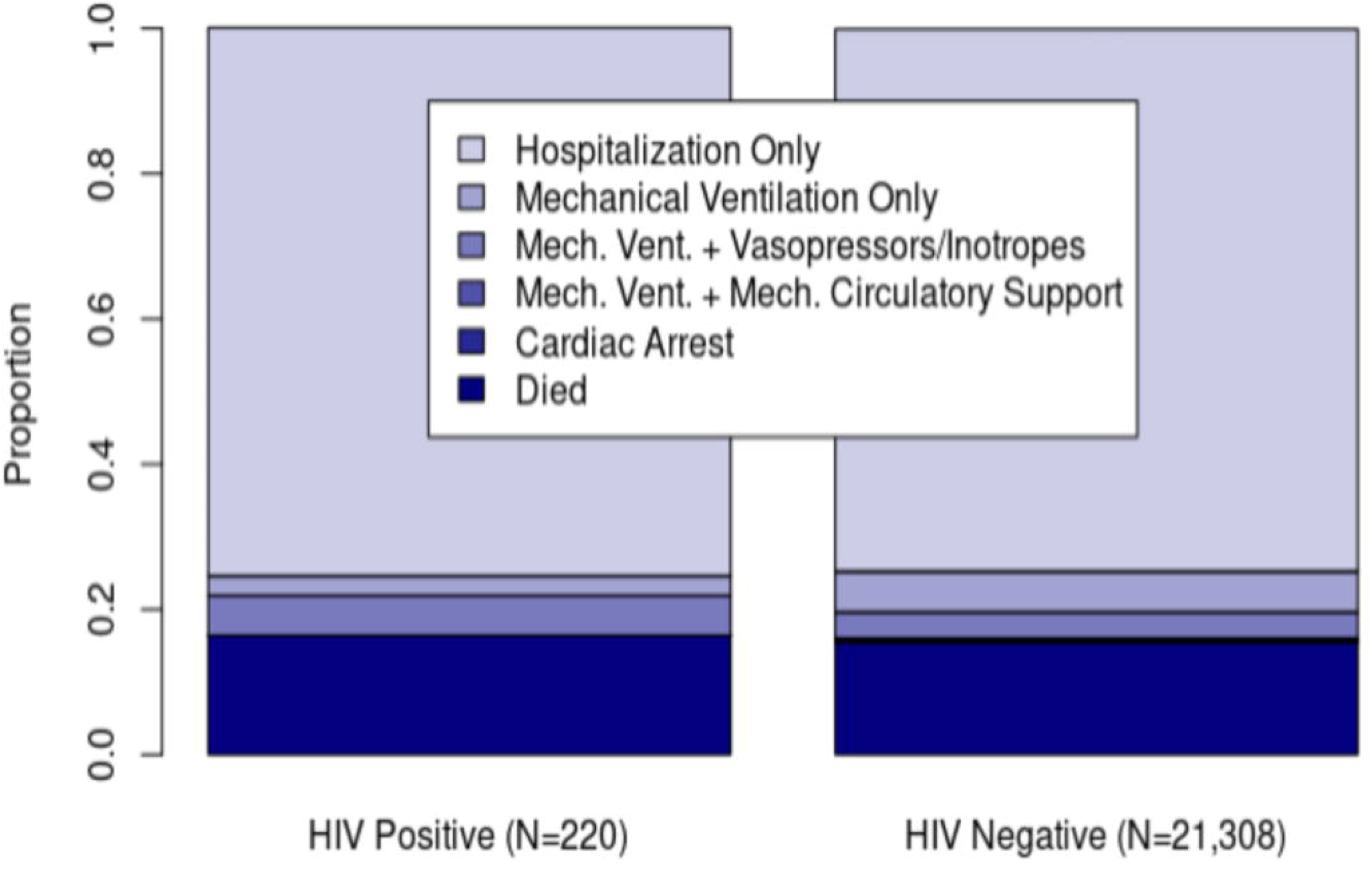
Severity of Illness by AHA COVID-19 Ordinal Status by HIV. HIV is not associated with differences in clinical severity as defined by the AHA COVID Ordinal Scale: Hospitalization, Mechanical Ventilation, Mechanical Ventilation and Vasopressors/Inotropes, Mechanical Ventilation and Mechanical Circulatory Support, Cardiac Arrest, and Death; (p=0.93 by Kruskal-Wallis test) which persisted with adjustment for clustering by hospital and patient characteristics (aOR 0.96, 95%CI 0.62-1.50, p=0.86). Among those with HIV, 47 (21.4%) required endotracheal intubation and mechanical ventilation compared to 4,152 (19.3%) without HIV (aOR 1.00; 95%CI. 0.71-1.40; p=1.00). Similarly, 59 PLWH (26.8%) compared to 6,604 without HIV (30.7%) were managed in an intensive care unit (aOR 0.90; 95%CI 0.65-1.23; p=0.49).

HIV was also not associated with differences in length of stay by unadjusted analysis (median 6 days with or without HIV; p=0.73). After adjustment for demographics and past medical history, HIV was not associated with an increased length of stay (aOR 1.03; 95% CI 0.76-1.66; p=0.21). Overall participation in a COVID-19 clinical trial was low and did not differ by HIV status (8.6% among PLWH and 9.2% among people without HIV, p=0.80).

As a sensitivity analysis using propensity matching as an alternative approach to account for confounding, we found an increased effect size compatible with an association of HIV with increased risk of in-hospital mortality that was not statistically significant (OR 1.53; 95% CI 0.96-2.42; p=0.06). In the propensity-matched analysis, there was not a significant association between HIV and MACE (aOR 1.17; 95%CI 0.76-1.77; p=0.45).

## Discussion

Among those hospitalized for COVID-19 at 107 hospitals participating in the AHA COVID-19 Cardiovascular Disease registry, PLWH were younger and more likely to identify as Non-Hispanic Black, consistent with the demographics of PLWH in the United States. We found that HIV was not associated with in-hospital mortality among people hospitalized with COVID-19 in both unadjusted and adjusted models. We also did not find significant differences in major adverse cardiac events, severity of illness including intensive care unit admission and mechanical ventilation, or length of stay by HIV status. Lastly, enrollment in COVID-19 clinical trials was not lower among PLWH compared to people without HIV.

### Hypotheses regarding HIV as a risk factor

The biologic rationale underlying the impact of HIV infection on COVID-19 outcomes could include worsening of clinical sequelae or a paradoxical protective effect. Namely, HIV could either decrease risk of mortality among PLWH via reduced severity of illness either by decreased immune response or partial effectiveness of HIV-targeted antiretroviral therapy against SARS-CoV-2, or increase risk of mortality due to changes in coagulation or increased cardiovascular risk factors. In our study, the severity of illness among HIV-infected individuals hospitalized with COVID-19 was similar to individuals without HIV, and inflammatory biomarkers were similar. Despite having higher d-dimer levels and higher rates of traditional cardiovascular risk factors, PLWH had similar rates of MACE and troponin levels compared to uninfected individuals.

### Comparison with other studies of hospitalized PLWH

As the largest hospital-based analysis in the United States to date, our finding that HIV is not independently associated with in-hospital mortality is consistent with most prior reports among PLWH hospitalized with COVID-19 (10-15). Also consistent with our results, the Veterans Aging Cohort Study, which included 253 HIV+/COVID+ veterans, most of whom were not hospitalized, found similar risk of hospital admission (aHR 1.09, 95% CI 0.85 to 1.41), intensive care unit admission (aHR 1.08, 95% CI 0.72 to 1.62), and death (aHR 1.08, 95% CI 0.66 to 1.75) (14). Our study has additional power to confirm these findings among hospitalized patients.

Prior to our study, the two largest studies of PLWH hospitalized for COVID-19 were the HIV-COVID-19 consortium study (21) and the United Kingdom-based ISARIC WHO CCP study, which included 122 PLWH hospitalized for COVID-19 and an HIV-negative control group (16). The HIV-COVID-19 consortium study was a multicenter registry that included 286 PLWH at 36 institutions mostly in the United States, of whom 164 were hospitalized. This descriptive study lacked a control group of people without HIV, but found a mortality rate or 16.5%, similar to our study. The ISARIC WHO CCP investigators found higher overall mortality than our study or the consortium study (26.7% among PLWH, 32.1% among those without HIV). They found no difference in crude mortality by HIV status, which is consistent with our findings. After adjustment for age, however, they found that HIV was associated with higher risk of mortality (HR 1.47, 95% CI 1.01-2.14). Similar to their analyses, our results did not change despite adjusting for comorbidities. Our data provide evidence that PLWH hospitalized with COVID-19 in the United States are not at significantly elevated risk of complications from COVID-19 infection including in-hospital mortality, intensive care unit admission, need for mechanical ventilation, major adverse cardiovascular events, or prolonged length of stay.

### Distinction of our findings from population-based registry data

Our results among hospitalized PLWH contrast with findings of increased risk found in some population-based registry studies. In particular, a large population-based registry in the United Kingdom (OpenSAFELY) (22) and a South African cohort study (7) both found higher population-based mortality rates among PLWH with COVID-19 compared to those without PLWH. A population-based approach may yield different results if HIV status is associated with differential rates of admission overall, as shown in some studies (6,9), or presentation to a participating hospital. In contrast to those studies, we estimated the association of HIV infection with in-hospital mortality conditional upon being admitted to a participating hospital as opposed to calculating the marginal risk across a population. Therefore, the results of our hospital-based analysis may be consistent with a meaningfully increased population-level risk among PLWH because our analysis is restricted to those with severe disease requiring hospitalization (22). Future studies may provide insights reconciling population-level differences in outcomes with our findings, which may be related to social determinants of health, differences in infection rates, and differences in severity (17).

### Limitations

The strengths of our study are that this is a large study population with over 21,000 people hospitalized at 107 hospitals with rigorous clinical adjudication of outcomes and detailed clinical phenotyping. The registry did not contain details regarding HIV such as treatment with antiretroviral therapy, CD4 counts, or viral loads. Despite diversity in hospital type and geographic location, the subset of hospitals participating in the registry may not be representative of all hospitals in the United States. Therefore, we focused not a population-based analysis to estimate marginal effects of the average additional risk of HIV but rather the conditional effect: what additional risk might a person living with HIV face conditional upon being admitted to a participating hospital. While the absolute number of HIV-infected individuals with MACE and death was low, our study does represent the largest study of PLWH hospitalized with COVID-19 to date. As an observational study, it remains possible that there is unmeasured confounding; to address this concern, we performed a propensity score-matched analysis in which we achieved good matching, but the results of this analysis were compatible with our findings as well as a significant association. As data are entered by each individual site, it is possible that there are variations in interpretation of the case definitions. We also did not account for calendar week or the local burden of the pandemic at a given hospital site as we hypothesized these should be non-differential with respect to HIV status.

## Conclusions

In this registry-based study of over 21,000 people hospitalized for COVID-19, 1.1% had a past medical history that included HIV. Despite having more traditional cardiovascular risk factors, HIV is not independently associated with increased incidence of in-hospital mortality, incidence of MACE, or COVID-19 severity among adults hospitalized for COVID-19 at registry-participating hospitals. Future studies evaluating the long-term sequelae of COVID-19 in the setting of HIV infection are needed.

## Perspectives

### Core Clinical Competencies

#### Competency in Patient Care

Despite having more cardiovascular risk factors, HIV individuals hospitalized for COVID 19 did not have worsened in-hospital mortality, ordinal score for COVID-19 or MACE compared to non-HIV infected individuals.

#### Translational Outlook

Additional studies are needed to ascertain the clinical ramifications of HIV infection and COVID-19 infection. While among individuals hospitalized for COVID 9, PLWH appear to be similar to uninfected individuals, the impact of HIV infection, chronic inflammation and immune activation and long-term sequelae of COVID 19 in the setting of HIV remains unknown.

## Data Availability

De-identified data from the American Heart Association's COVID-19 registry is available through the AHA's Precision Medicine Platform. Requests to access the data should be made through the AHA COVID registry after submitting a proposal:
https://www.heart.org/en/professional/quality-improvement/covid-19-cvd-registry/covid-19-cvdregistry-research-opportunities.

## Acknowledgements

The Get With The Guidelines® programs are provided by the American Heart Association (AHA). The AHA Precision Medicine Platform (https://precision.heart.org/) was used for data analysis. IQVIA (Parsippany, New Jersey) serves as the data collection and coordination center.

## Abbreviations

(HIV): human immunodeficiency virus
(PLWH): people living with HIV
(MACE): major adverse cardiac events
(aOR): adjusted odds ratio
(LOS): length of stay
(AHA): American Heart Association
(AIDS): acquired immunodeficiency syndrome
(BMI): body mass index
(COVID-19): coronavirus disease 2019

## Notes

**Sources of funding:** AHA’s suite of Registries is funded by multiple industry sponsors. AHA’s COVID-19 CVD Registry is partially supported by The Gordon and Betty Moore Foundation (Palo Alto, California). Dr Durstenfeld is supported by NIH/NHLBI 5T32HL007731-28. Dr. Rodriguez was funded by NIH/NHLBI K01HL144607 and AHA/Robert Wood Johnson Harold Amos Medical Faculty Development Program. Dr Secemsky is supported by NIH/NHLBI K23HL150290. Dr. Parikh is supported by AHA 18CDA34110335. Dr Hsue is supported by NIH/NIAID 2K24AI112393-06.

### Competing Interest Statement

Disclosures: MSD, KS, and YM have no disclosures. FR has received consulting fees from Novartis, Janssen, NovoNordisk, and HealthPals unrelated to this work. EAS reports unrelated research grants to BIDMC: AstraZeneca, BD, Boston Scientific, Cook, CSI, Laminate Medical, Medtronic and Philips and unrelated consulting/speaking fees from Abbott, Bayer, BD, Boston Scientific, Cook, CSI, Inari, Janssen, Medtronic, Philips, and VentureMed. RVP reports unrelated research support Janssen and Infraredx; consulting fees from Abbott Vascular; and scientific advisory board (minor equity interest) of Stallion Cardio, DocVocate, and HeartCloud. PYH has received honoraria from Gilead and Merck, research grant from Novartis, unrelated to this work..

### Funding Statement

Sources of funding: The authors did not receive payment for any aspects of the submitted work. AHA's suite of Registries is funded by multiple industry sponsors. AHA's COVID-19 CVD Registry is partially supported by The Gordon and Betty Moore Foundation (Palo Alto, California). Dr. Durstenfeld is supported by NIH/NHLBI 5T32HL007731-28. Dr. Rodriguez is funded by NIH/NHLBI K01 HL 144607 and the American Heart Association/Robert Wood Johnson Harold Amos Medical Faculty Development Program. Dr. Secemsky is supported by NIH/NHLBI K23HL150290. Dr. Parikh is supported by AHA grant 18CDA34110335. Dr. Hsue is supported by NIH/NIAID 2K24AI112393-06.

### Author Declarations

University of California, San Francisco provided IRB approval for this study. A waiver of consent was granted and the analysis was conducted with a deidentified dataset.

### Summary of Updates

Additional endpoints added to text and Table 2.

