## Supplemental Appendix for "Impact of HIV Infection on COVID-19 Outcomes Among Hospitalized Adults in the U.S."

**Supplemental Appendix 1: Past medical history by HIV status**

| **Past Medical History** | **HIV positive (n=220)** | **HIV negative (n=21,308)** | **p-value** |
| --- | --- | --- | --- |
| Atrial Fibrillation | 12 (5.5%) | 2001 (9.3%) | 0.06 |
| Atrial Flutter | 2 (0.9%) | 152 (0.7%) | 0.67 |
| Cancer | 27 (12.3%) | 2680 (12.4%) | 1 |
| Cerebrovascular Disease | 27 (12.3%) | 2608 (12.1%) | 1 |
| Stroke | 24 (10.9%) | 2060 (9.6%) | 0.57 |
| Transient Ischemic Attack | 5 (2.3%) | 625 (2.9%) | 0.72 |
| Currently on Dialysis | 13 (5.9%) | 741 (3.4%) | 0.07 |
| Diabetes Mellitus | 88 (40.0%) | 7614 (35.4%) | 0.17 |
| Dyslipidemia | 69 (31.4% | 7423 (34.5%) | 0.37 |
| Heart Failure | 24 (10.9%) | 2500 (11.6%) | 0.83 |
| Hypertension | 125 (56.8%) | 12673 (58.9%) | 0.58 |
| Peripheral Artery Disease | 3 (1.4%) | 584 (2.7%) | 0.30 |
| Prior CABG | 2 (0.9%) | 627 (2.9%) | 0.10 |
| Prior MI | 9 (4.1%) | 1199 (5.6%) | 0.42 |
| Prior PCI | 6 (2.7%) | 986 (4.6%) | 0.25 |
| No Medical History | 0 | 3792 (17.6%) | <0.001 |
| Chronic Kidney Disease | 35 (15.9%) | 2784 (12.9%) | 0.22 |
| DVT | 7 (3.2%) | 725 (3.4%) | 1 |
| eCigarette (vaping) | 0 | 31 (0.1%) | 1 |
| Smoking | 28 (12.7%) | 1406 (6.5%) | <0.001 |
| Lupus | 0 | 121 (0.6%) | 0.64 |
| Rheumatoid Arthritis | 4 (1.8%) | 242 (1.1%) | 0.32 |
| Other Immune Disorder | 3 (1.4%) | 411 (1.9%) | 0.80 |
| Pulmonary Embolism | 4 (1.8%) | 487 (2.3%) | 0.82 |
| Pulmonary Disease | 48 (21.8%) | 4015 (18.7) | 0.26 |
| COPD | 24 (10.9%) | 1879 (8.7%) | 0.30 |
| Interstitial Lung Disease (ILD) | 0 | 100 (0.5%) | 0.63 |
| Asthma | 25 (11.4%) | 1948 (9.0%) | 0.28 |
| Other Pulmonary Disease | 5 (2.3%) | 535 (2.5%) | 1 |
| Organ Transplant | 2 (0.9%) | 370 (1.7%) | 0.60 |
| Congenital heart disease | 0 | 58 (0.3%) | 1 |
| Pulmonary Arterial Hypertension | 2 (0.9%) | 106 (0.5%) | 0.30 |
